# Comparison of Gastric Alimetry^®^ body surface gastric mapping versus electrogastrography spectral analysis

**DOI:** 10.1101/2023.06.05.23290993

**Authors:** Gabriel Schamberg, Stefan Calder, Chris Varghese, William Xu, William Jiaen Wang, Vincent Ho, Charlotte Daker, Christopher N. Andrews, Greg O’Grady, Armen A. Gharibans

## Abstract

Electrogastrography (EGG) non-invasively evaluates gastric motility but is viewed as lacking clinical utility. Gastric Alimetry® is a new diagnostic test that combines high-resolution body surface gastric mapping (BSGM) with validated symptom profiling, with the goal of overcoming EGG’s limitations. This study directly compared EGG and BSGM to define performance differences in spectral analysis. Comparisons between Gastric Alimetry BSGM and EGG were conducted by protocolized evaluation of 178 subjects (110 controls; 68 nausea and vomiting (NVS) and/or type 1 diabetes (T1D)). Comparisons followed standard methodologies for each test (pre-processing, post-processing, analysis), with statistical evaluations for group-level differences, symptom correlations, and patient-level classifications. BSGM showed substantially tighter frequency ranges vs EGG in controls. Both tests detected rhythm instability in NVS, but EGG showed opposite frequency effects in T1D. BSGM showed an 8x increase in the number of significant correlations with symptoms. BSGM accuracy for patient-level classification was 0.78 for patients v. controls and 0.96 as compared to blinded consensus panel; EGG accuracy was 0.54 and 0.43. EGG detected group-level differences in patients, but lacked symptom correlations and showed poor accuracy for patient-level classification, explaining EGG’s limited clinical utility. BSGM demonstrated substantial performance improvements across all domains.

## Introduction

Chronic gastroduodenal symptoms affect >10% of the population and impart a vast health burden, however, their diagnosis and management remains challenging.^1^ Current symptom-based classifications overlap, and gastric emptying testing has recently been shown to be labile over time and insensitive for gastric neuromuscular disorders.^2^ Improved diagnostic tests are needed to phenotype specific patient subgroups and inform personalized care.^3, 4^

Electrogastrography (EGG) has been applied to non-invasively evaluate gastric function.^5–7^ A substantial EGG literature has accumulated over previous decades showing gastric myoelectrical abnormalities are prevalent in gastroduodenal disorders, including chronic nausea and vomiting, gastroparesis, functional dyspepsia, and reflux disease.^8–11^ However, as a clinical tool, EGG has achieved limited impact owing to multiple technical limitations, including high sensitivity to noise and questionable ability to reliably identify specific patient populations.^3, 12–14^

Gastric Alimetry® (Alimetry, New Zealand) is a new test of gastric function involving simultaneous body surface gastric mapping (BSGM) and validated symptom profiling.^15–17^ Many technical improvements have been implemented that together offer a fundamental advance over traditional EGG.^18^ These include conformable electronics for high-resolution spatial sampling over a dense field (8×8 electrodes; 256 cm^2^), patient-specific positioning to counter gastric anatomical variability, wearable hardware to minimize motion artifacts, and a validated signal processing pipeline that maximally separates weak gastric signals from noise.^16, 17, 19, 20^ Further advances include a standardized test methodology,^17^ new spectral metrics that resolve pitfalls in traditional EGG analytics,^21^ and reference intervals for the new metrics developed on a large and diverse cohort of healthy volunteers.^22^ These techniques have recently been applied in clinical studies to achieve patient phenotyping and symptom correlations in nausea and vomiting disorders, functional dyspepsia and type 1 diabetes (T1D).^17, 23, 24^

This study aimed to compare Gastric Alimetry BSGM and EGG spectral metrics in a highly standardized manner to quantify performance differences. This comparison was conducted on parallel end-to-end BSGM and EGG analytical pipelines in order to assess the collective impact of the improvements in test protocol, hardware, signal processing, and analysis metrics developed for Gastric Alimetry BSGM. Specifically, we assessed the relative utilities of BSGM and EGG in three domains: i) establishing group-level differences in measures of gastric activity; ii) capturing the relationship between gastric abnormalities and symptoms; and iii) yielding patient-level classifications of gastric health.

## Methods

### Study population

Head to head comparisons between BSGM and EGG were conducted by protocolised evaluation of an existing database of 178 subjects, comprising 110 healthy volunteers, 32 patients with type 1 diabetes (T1D), and 43 patients with chronic nausea and vomiting syndromes (NVS) (7 patients had both T1D and NVS).^172223^ Data collection was conducted in Auckland (New Zealand), Calgary (Canada), Louisville (KY, United States), and Western Sydney (Australia). The study protocols were approved by The Auckland Health Research Ethics Committee (AHREC; AH1130), The University of Calgary Conjoint Health Research Ethics Board (REB19-1925), University of Louisville IRB (Ref 723369), and the Human Research Ethics Committee at Western Sydney (H13541). All data was collected in accordance with the guidelines and regulations of these committees. All subjects provided informed consent.

Comparisons were performed according to standard methodological and analysis pipelines for each test, as outlined in **Figure 1** and described in the sections below. To ensure a fair comparison, all implemented methods and criteria within both pipelines were referenced against standardized techniques in the literature, without deviation from commonly accepted practices, with a fully-automated computational analysis.

**Figure 1.**
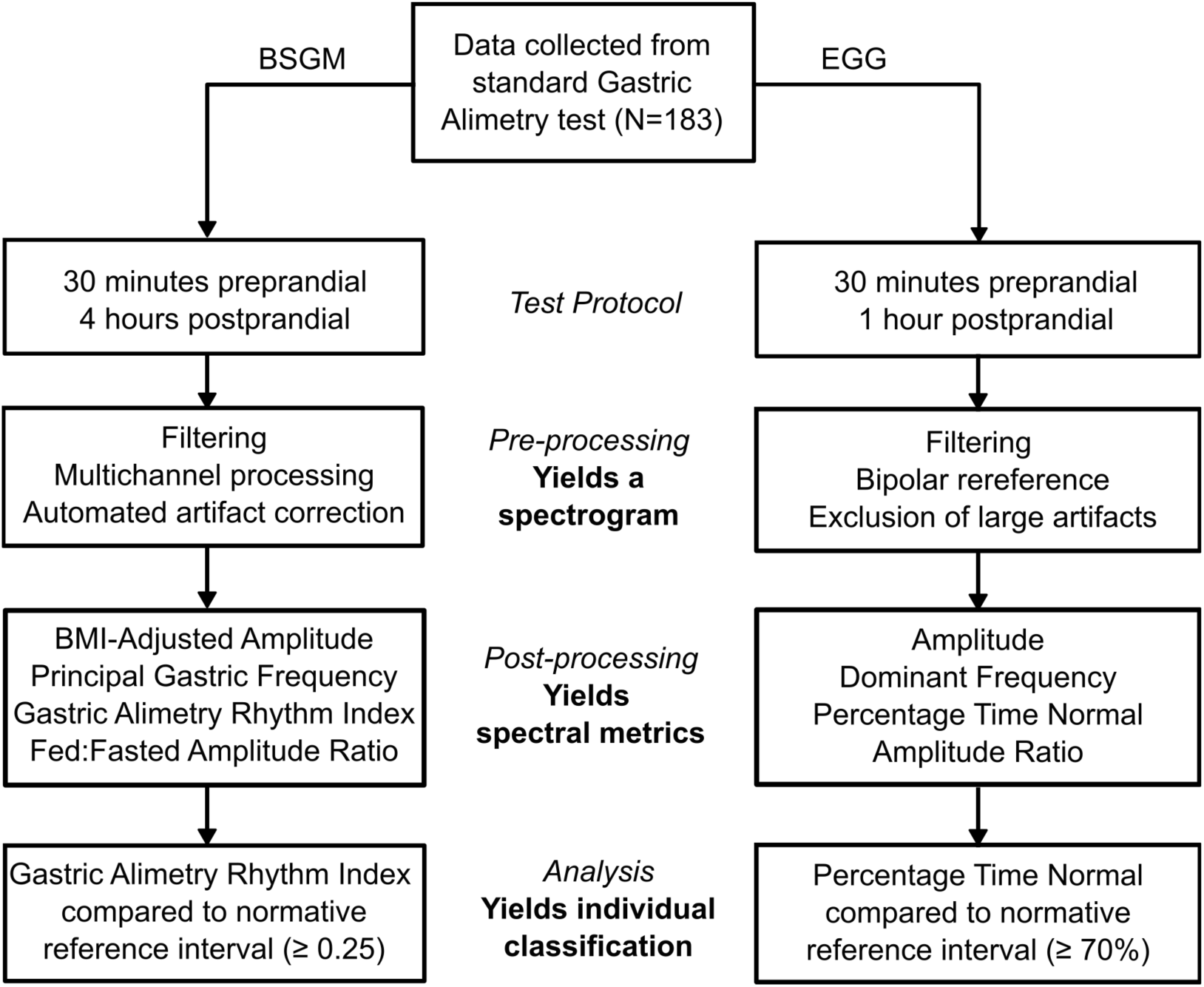
Flow chart depicting the relevant differences between the BSGM and EGG pipelines at the data collection, pre-processing, post-processing, and analysis stages.

### Test protocol

Data collection for the EGG and BSGM pipelines was identical. All subjects fasted for >6 hrs and avoided caffeine and nicotine prior to testing. Medications known to influence gut motility were withheld for 72 hrs prior to testing. Electrode array placement was preceded by skin preparation (NuPrep; Weaver & Co, CO, USA), with impedance checks prior to commencing recordings. Data was collected over a fasting period of 30 minutes, followed by a 482 kCal meal consumed over 10 minutes, followed by a 4-hr postprandial recording. The meal consisted of an oatmeal energy bar (250 kcal, 5 g fat, 45 g carbohydrate, 10 g protein, 7 g fiber; Clif Bar & Company, CA, USA) and Ensure (232 kcal, 250 mL; Abbott Nutrition, IL, USA), or an appropriate calorie-matched diabetic meal substitute.^23^ Participants sat reclined in a chair and were asked to limit movement, talking, and sleeping, but were able to read, watch media, work on a mobile device, and mobilize for comfort breaks. Throughout the test, patients logged their symptoms on an iPad using a validated app.^15^ When analyzing the data using the EGG pipeline, only the first 90 minutes of the recorded data was used (30 minutes fasted, 60 minutes fed). A one-hour postprandial period was chosen as this is at the upper end of the suggested durations in the EGG literature.^7, 25^

### Pre-processing

All data was collected using the electrode array and hardware of the Gastric Alimetry system, which were then processed via the separate BSGM and EGG analytical pipelines in parallel. Data preprocessing in the BSGM pipeline was performed using the Gastric Alimetry algorithm.^16, 17^ The steps in the algorithm include filtering, automated identification and correction of motion artifacts, and multichannel processing. The result of these steps is a single spectrogram, summated from the top-ranked channels of the Gastric Alimetry array, which describes the frequency and amplitude content of recorded signals as a function of time over the course of the 4.5 hour test.^16^

The EGG preprocessing pipeline was designed to most closely match standardized techniques established in the literature.^7, 14, 25–27^ All data was bandpass filtered to the 1-6 cpm frequency band. Two channels from the complete 64-channel array that most closely represented the recommended optimal placement of the EGG electrodes were selected (**Figure 2**). Specifically, we identified the electrode along the midline closest to the midpoint between the umbilicus and the xiphoid, along with a reference electrode at equal height and 6 cm to the right of the main electrode.^26^ If either of the selected electrodes had poor skin contact based on an automated impedance check of all channels,^16^ then the next closest pair was identified. This process was repeated until a pair of electrodes with sufficient skin contact was identified (see **Figure 2**). The data was transformed using a bipolar reference, yielding a single EGG recording at the optimal specified location, consistent with an estimated location overlying the gastric antrum.^28^

**Figure 2.**
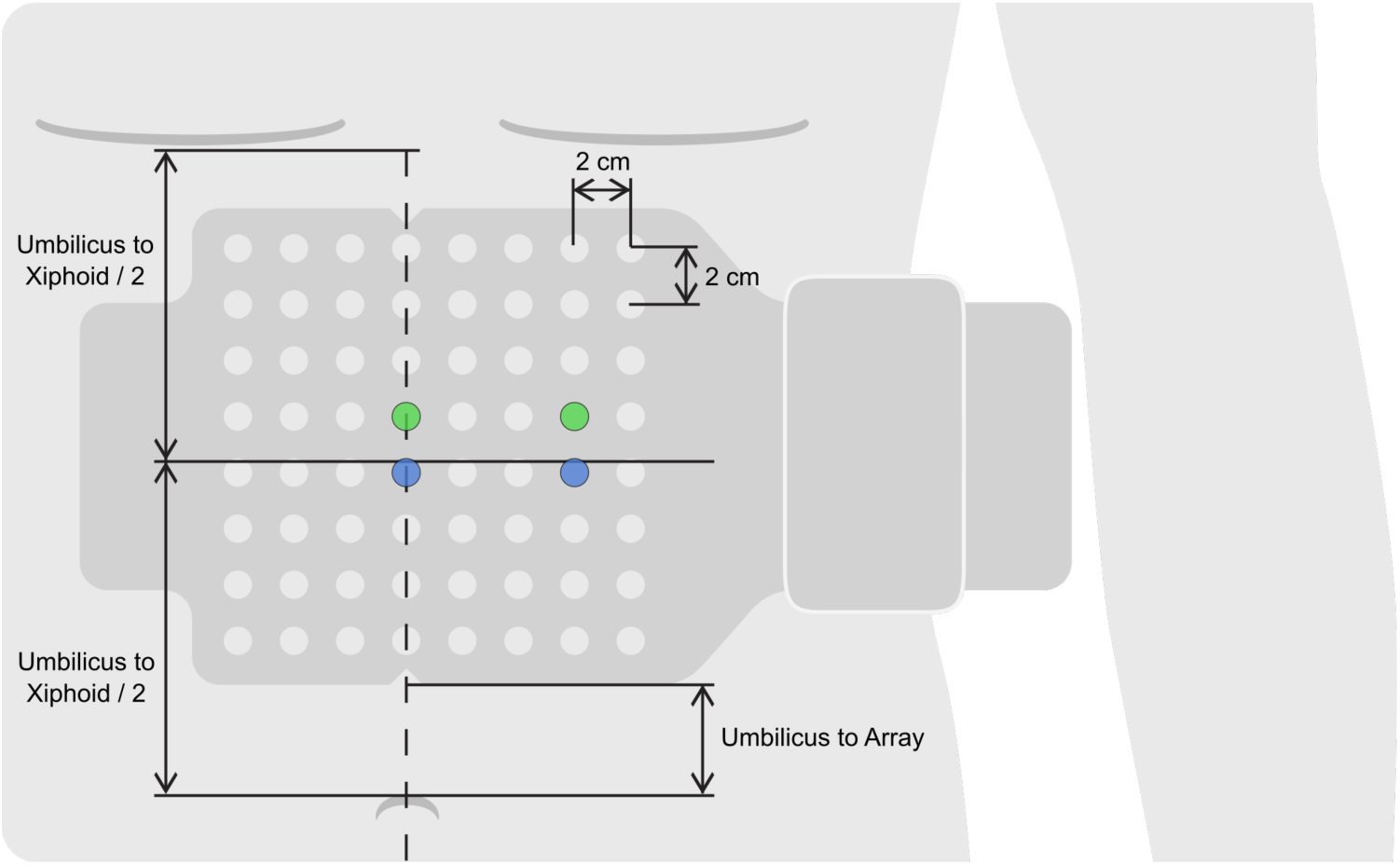
BSGM array placement and EGG electrode identification procedure. The main electrode was selected as the electrode along the dotted line closest to the midpoint between the umbilicus and xiphoid (blue, left). The reference electrode was selected as the electrode three to the right in the same row as the main electrode (blue, right). If either of those electrodes had impedance > 500 kilohms, the next closest electrodes were selected (green)

Commercially available EGG devices do not perform any artifact correction, but they do enable marking of time periods with artifacts for removal from further processing, either by hand or using proprietary artifact detection algorithms. As manual marking of artifacts is not practical for the large dataset analyzed in this study, we used a recently-developed artifact detection algorithm validated against expert manual marking to mark time periods for removal in the EGG pipeline.^19, 29^ The resulting EGG signal with artifacts removed was used to generate a spectrogram per Gharibans et al.^16^

### Post-processing

The respective spectrograms were then used to compute a collection of spectral metrics typically used in BSGM and EGG analyses. For each pipeline, four metrics were computed for the purpose of characterizing the strength, frequency, rhythmic stability, and responsiveness to meal stimulus of the recorded gastric activity, respectively given by:

- ***Amplitude*** *(EGG)* - average of the amplitude over the 90 minute recording
- ***BMI-Adjusted Amplitude*** *(BSGM) -* average whole-test amplitude adjusted for BMI as a proxy for adipose tissue
- ***Dominant Frequency*** *(EGG) -* frequency associated with the highest amplitude in the overall average spectrum
- ***Principal Gastric Frequency*** *(BSGM)* - Frequency associated with highest Gastric Alimetry Rhythm Index
- ***Percentage Time Normal Frequency*** *(EGG)* - percentage of (non-artifact) windows where the frequency with maximal amplitude is between 2-4 cpm
- ***Gastric Alimetry Rhythm Index (GA-RI)^TM^*** *(BSGM)* - Measure of the concentration of amplitude in a narrow frequency band over time
- ***Amplitude Ratio*** *(EGG) -* Average postprandial amplitude divided by average preprandial amplitude
- ***Fed:Fasted Amplitude Ratio (ff-AR)*** *(BSGM) -* Maximal 1-hour average postprandial amplitude divided by average preprandial amplitude

Further details for the EGG and BSGM metrics are available in Yin et al. and Chang 2005 ^7, 27^, and Varghese et al. and Schamberg et al.^21, 22^, respectively. In some instances, one or more BSGM metrics may not be reported (for example, no Principal Gastric Frequency is reported in severely dysrhythmic cases). While some EGG studies exclude metrics in some instances (e.g., when a dominant frequency is not prominent), the details on how these calculations are made are not publicly available and therefore were not implemented in this study.

### Statistical Analyses

#### i) Group-Level Differences in Spectral Metrics

The eight metrics (four BSGM, four EGG) were calculated for all subjects and reported as median and interquartile range (IQR) for the control, T1D, and NVS subgroups. For each metric, an independent t-test was performed to test for group-level differences between the control group and each patient subgroup. As the amplitude and amplitude ratio metrics were highly skewed (for BSGM and EGG), these variables were log transformed before performing statistical analyses. BSGM metrics that are not reported are excluded from the t-test and reported separately.

#### ii) Correlation of Spectral Metrics with Symptoms

The Pearson correlation coefficient with 95% bootstrap confidence interval and associated p-value were calculated for each metric/symptom pair on the entire cohort. The symptom values were calculated as the whole-test average for each symptom recorded by the patients in the symptom logging App (upper gut pain, nausea, bloating, heartburn, stomach burn, and excessive fullness), along with the Total Symptom Burden Score, calculated as the sum of postprandial averages of each symptom plus the patient’s reported early satiation.^15^ To account for the large number of correlations tested (eight metrics, seven symptoms, 56 total correlations), significance was assessed using the Benjamini-Hochberg correction to control for a 10% false discovery rate.^30^ Given that frequency abnormalities can occur at both high and low frequencies, correlations between symptoms and frequency metrics were computed using the absolute difference of the frequency metric from 3 cpm (termed ‘frequency deviation’).

#### iii) Individual-Level Classification of NVS Patients and Healthy Controls (Phenotyping)

It has been demonstrated in the literature that NVS is associated with dysrhythmic gastric activity in a subgroup of affected patients.^10, 17, 31^ Using published reference values for measures of rhythmic stability (0.25 for GA-RI in BSGM ^22^; and 70% for percentage time normal frequency in EGG ^7, 14, 28, 32– 34^), each of the controls and NVS patients was classified as having normal or abnormal rhythmic stability for each test. These classifications were compared against each subject’s true status of NVS patient (n=43) or control (n=100). Given that a significant subgroup of NVS patients are known to have normal gastric activity^17^, it is expected that some NVS patients will not be identifiable from BSGM/EGG spectral analyses. As such, we additionally evaluated the classifications using the reference standard of NVS-Normal and NVS-Abnormal labels determined by an expert consensus classification panel who were blinded to patient or control status via a set of objective criteria (refer Gharibans et al ^17^). NVS patients labeled as ‘indeterminate’ by the consensus panel were excluded from this portion of the analysis (n=4). The classifications as determined by the EGG and BSGM metrics of rhythmic stability were compared with the subject subgroups of controls/NVS-Normal (n=136) and NVS-Abnormal (n=13) to generate overall diagnostic performance statistics. For both groupings, each subject was identified as a true positive (patient or NVS-Abnormal w/ abnormal stability; TP), true negative (control or control/NVS-Normal w/ normal stability; TN), false positive (control or control/NVS-Normal w/ abnormal stability; FP), or false negative (patient or NVS-Abnormal w/ normal stability; FN). These classifications were evaluated using sensitivity (TP/(TP+FN)), specificity (TN/(TN+FP)), and accuracy ((TP+TN)/(TP+TN+FP+FN)). Additionally, the utility of GA-RI and percentage time normal frequency for characterizing rhythmic abnormalities independent of the reference values was assessed using the area under the receiver operating characteristic (ROC) curve (AUC).

## Results

### Data Pre-and Post-processing

The procedure for selecting the bipolar pairs for use in EGG analysis selected the ideally located electrodes in 165 subjects (93%). The second closest electrode pair was used in 11 subjects and the third closest pair in one subject due to high impedance in the first and second closest pairs. For a single subject whose array positioning was not recorded, we used the central electrode referenced to an electrode 6 cm to the right.

The median (IQR) percentage of one-minute windows automatically excluded from the EGG spectrograms by the automated artifact detection algorithm was 36.5% (24.7% - 51.7%). Of the 178 cases analyzed with the BSGM pipeline, 54 spectrograms had windows excluded due to artifacts marked as unrecoverable by the Gastric Alimetry algorithm, with a maximum of 20% of windows excluded from a single case and a median (IQR) of 0% (0% - 0.7%).

All EGG metrics were reported for every subject. After analysis using the BSGM pipeline, 10 subjects had no identifiable Principal Gastric Frequency (two control, seven NVS, one T1D) and two subjects had no reported ff-AR due to >50% of the preprandial recording being removed due to high artifact content (one control, one NVS). All subjects had a reported GA-RI and BMI-Adjusted Amplitude. The median (IQR) for each metric, stratified by patient status, is provided in **Supplementary Table S1**.

Sample BSGM and EGG spectrograms for each of the three subject groups (control, T1D, and NVS) are shown in **Figure 3** and overall average spectrograms for each group are shown in **Supplementary Figure S1**.

**Figure 3.**
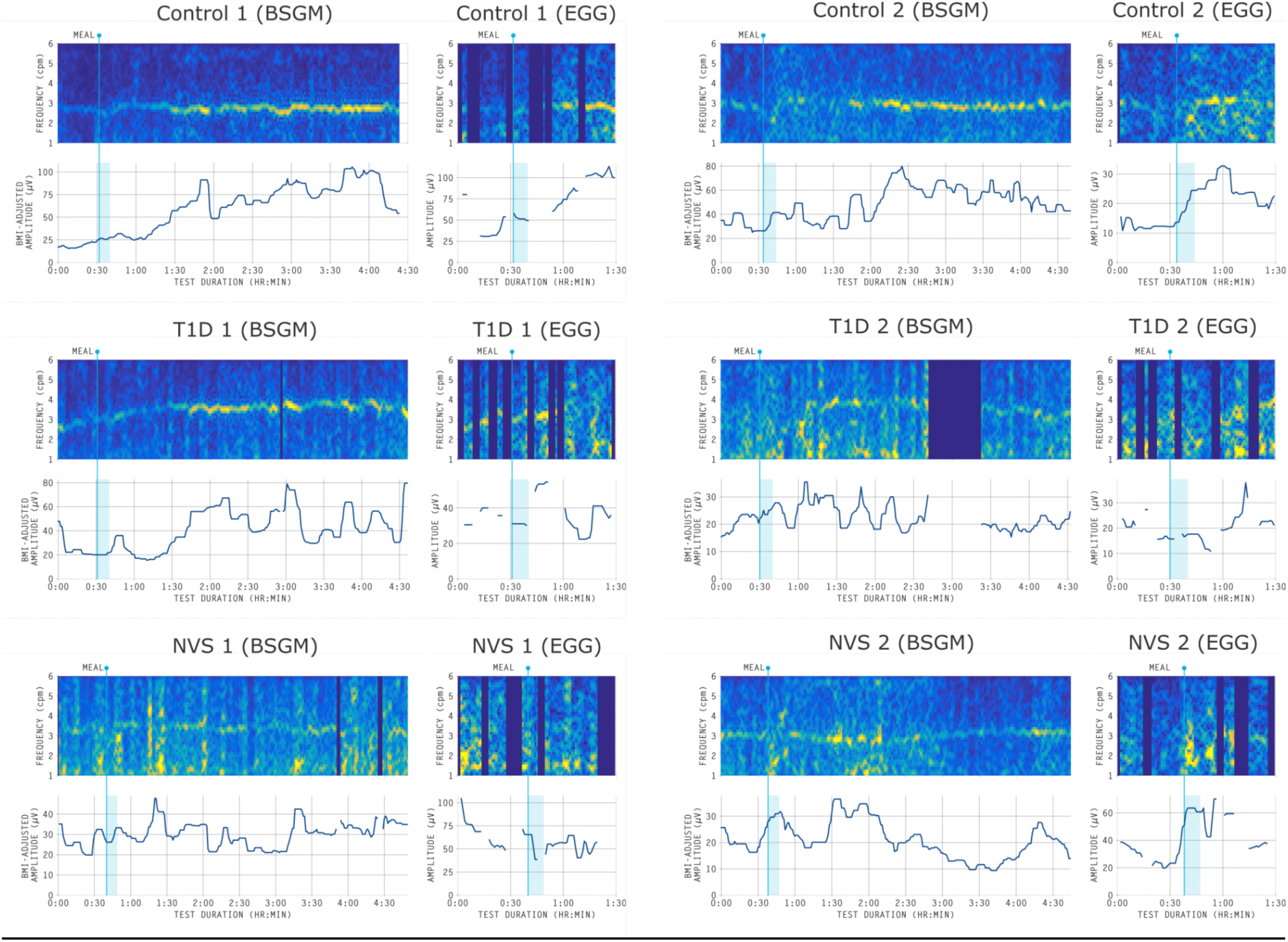
Representative BSGM and EGG spectrograms for two healthy controls, two T1D patients, and two NVS patients. Dark blue columns represent time periods marked as unrecoverable by the automated artifact algorithm in the BSGM spectrograms or marked as having an artifact in the EGG spectrograms.^19^ The BSGM spectrograms had very few time periods removed because the large majority of artifacts were corrected.

### Group-level differences

Boxplots of the group-level distributions for each BSGM and EGG metric are shown in **Figure 4**, with the median (IQR), t-statistic, and p-value for all metrics provided in **Supplementary Table S1**. It is known that the EGG dominant frequency metric has a pitfall of misidentifying low-frequency, high-amplitude transient activity as gastric activity, even in healthy controls.^2112^ This pitfall is illustrated by the marked difference in the BSGM Principal Gastric Frequency and EGG dominant frequency distributions among healthy controls (median 3.04 cpm (IQR 2.90 - 3.18) vs median 2.88 (IQR 1.50 - 3.12); p<0.0001). It has also recently been shown that healthy controls may exhibit considerable variability in the timing of peak gastric activity in response to meal consumption.^16, 21, 22^ This is observed in the improved performance of the BSGM vs EGG amplitude ratio metrics (median increase of 1.22x (IQR 1.00 - 1.46)). Notably, the longer test duration and optimized postprandial window selection used by the BSGM pipeline resulted in 17 controls having an EGG amplitude ratio <1 but a BSGM ff-AR >1.

**Figure 4.**
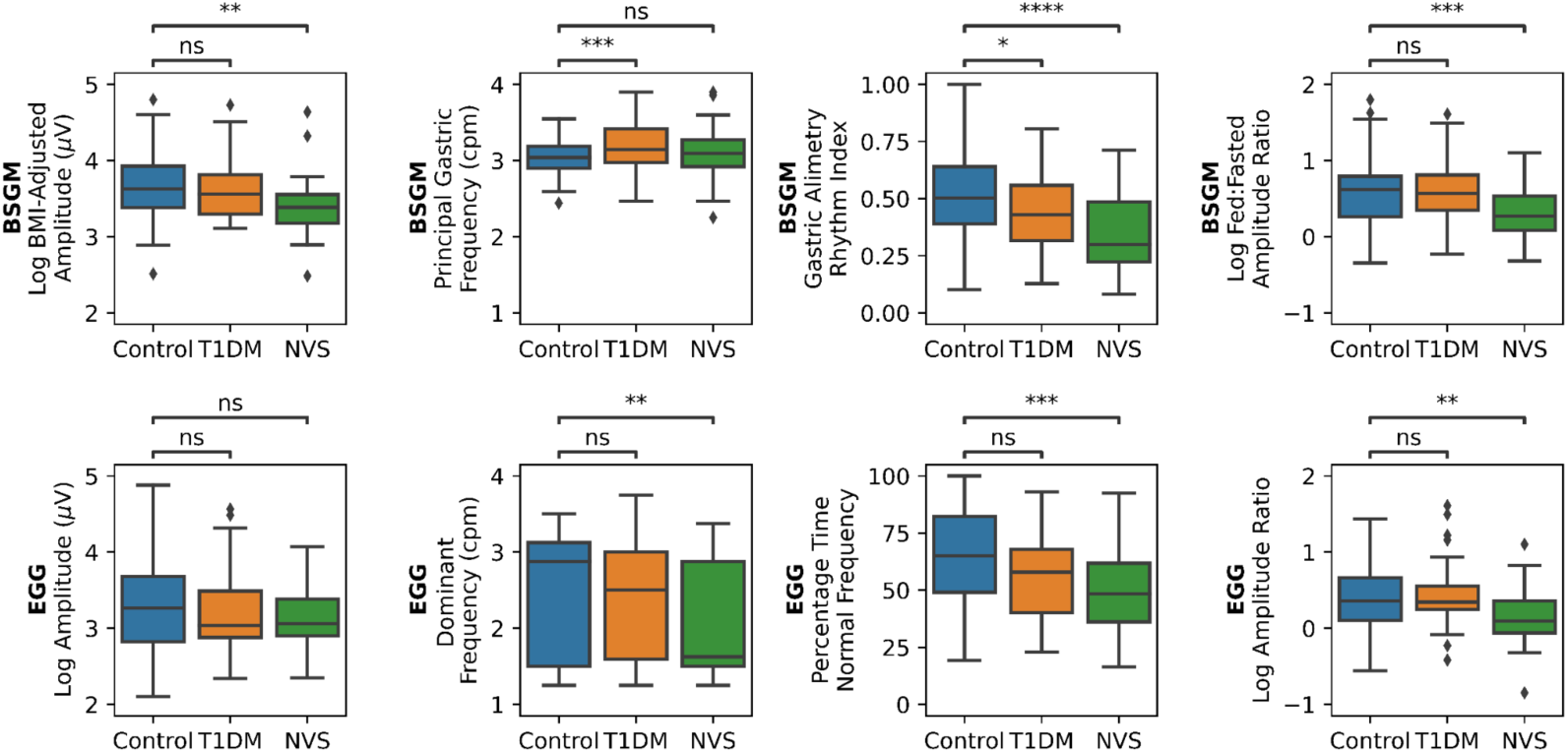
Group-level differences in metrics for BSGM (top) and EGG (bottom). Statistical significance is assessed with independent t-tests (ns: p>0.05, *: p<0.05, **: p<0.01, ***: p<0.001, ****: p<0.0001).

When comparing controls and T1D patients, two BSGM metrics were significantly different (Principal Gastric Frequency, p=0.0004; GA-RI, p=0.012) and one EGG metric was significantly different (percentage time normal frequency, p=0.045) (**Supplementary Table S1**). It can be observed in **Figure 4** that the lack of a significant difference between the EGG dominant frequency between controls and T1D patients is due to the large number of cases where the EGG dominant frequency is identified as a very low frequency (< 2 cpm), which is outside of the feasible frequency range for sustained gastric activity.^21, 35^ The EGG dominant frequency in T1D patients was decreased compared to controls (median 2.50 cpm vs. 2.88 cpm), which was the opposite effect to the BSGM Principal Gastric Frequency (median 3.15 cpm in T1D patients vs. 3.04 cpm in controls), which is notable because frequency deviation has recently been shown to be a correlate of symptoms in T1D patients by BSGM.^23^

When comparing controls and NVS patients, three BSGM metrics were significantly different (log BMI-Adjusted Amplitude, p=0.0005; GA-RI, p<0.0001; log ff-AR, p=0.0004) and three EGG metrics were significantly different (dominant frequency, p=0.0007; percentage time normal frequency, p=0.0003; log amplitude ratio p=0.0036). The difference in both BSGM and EGG rhythm stability metrics between controls and NVS patients is consistent with the well-described link between gastric dysrhythmia and NVS.^10, 17, 31, 36^ The absence of a difference in Principal Gastric Frequency reflects the ability of BSGM metrics to independently characterize gastric frequency and rhythm stability.^21^ Rather than identifying cases with no apparent coordinated gastric activity as having a low gastric frequency (as is the case with EGG dominant frequency), the BSGM pipeline instead did not report a Principal Gastric Frequency in seven of the NVS patients.

### Symptom Correlations

As shown in **Figure 5**, multiple BSGM metrics were correlated with patient symptoms logged during the test in the validated App (GA-RI with Total Symptom Burden, nausea, upper gut pain, and bloating; Principal Gastric Frequency deviation with excessive fullness, upper gut pain, and bloating; all p<0.05); whereas only the EGG percent time normal frequency metric and bloating symptom correlated (p=0.013). All correlations lay within the range of -0.3 < r < 0.3 reflecting the heterogeneous nature of the cohort. These low correlation coefficients are to be expected, as it is understood that a large subset of the patient population with high symptom scores will have normal gastric myoelectrical activity, reflecting alternative disease mechanisms.^37^. All significant correlation coefficients after adjustment for multiple comparison are shown in **Figure 5**, with the p-values, Benjamini-Hochberg critical values, and correlation coefficients with 95% bootstrap confidence intervals provided in **Supplementary Table S2.**

**Figure 5.**
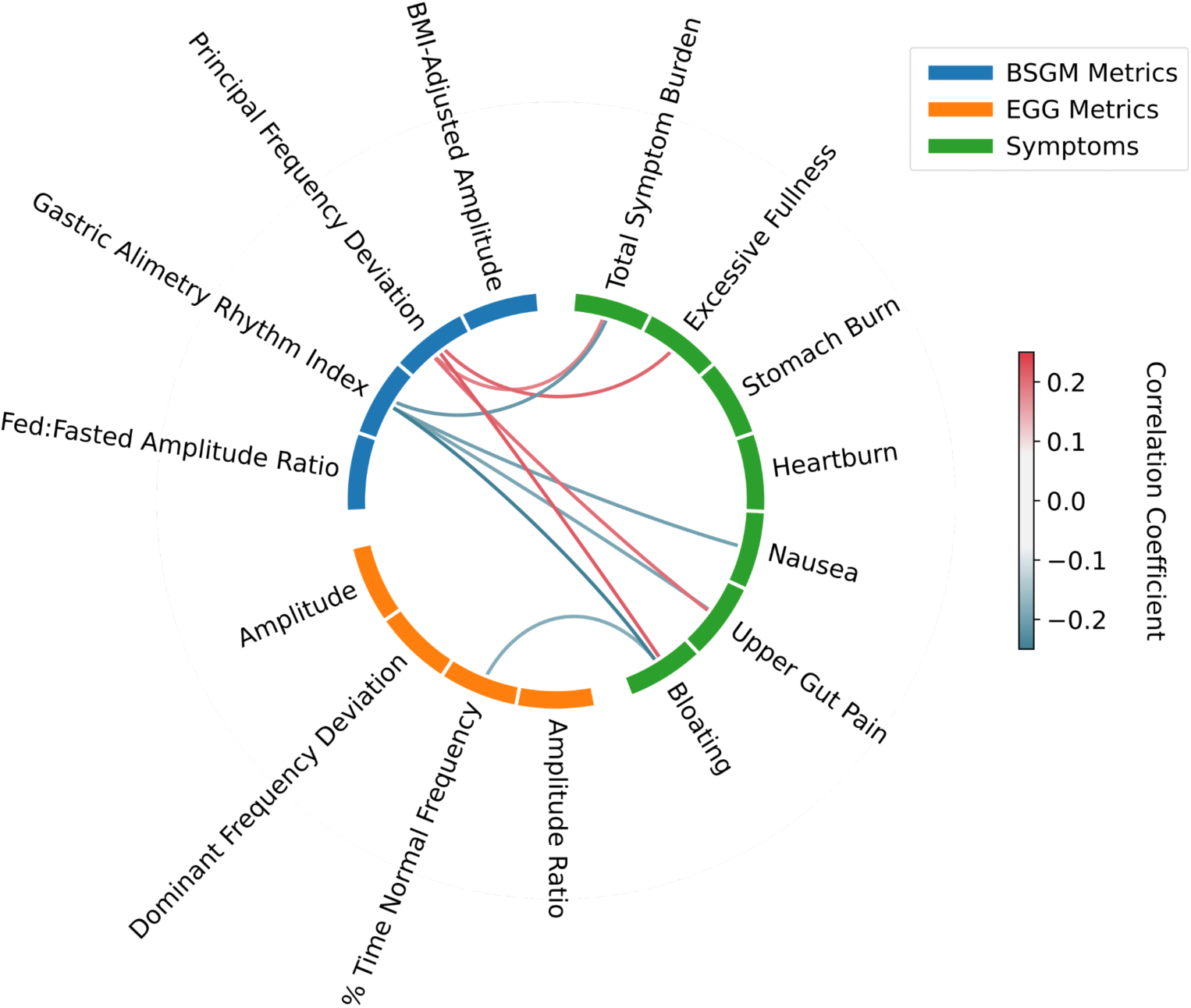
Correlations between metrics and symptoms. Only correlations that were significant after a Benjamini-Hochberg correction are shown. Only correlations between a metric and a symptom were tested. The range of measured correlations reflects the heterogeneous nature of the cohort, as it is understood that a large subset of the patient population with high symptom scores will have normal gastric myoelectrical activity and symptoms resulting from alternative disease mechanisms.

### Patient-Level Classification

The automated classifications of normal vs. abnormal gastric rhythm using BSGM and EGG, as compared to the two sets of ground truth labels, and associated ROC curves are shown in **Figure 6**. Quantitative performance metrics are provided in **Table 1**.

**Figure 6.**
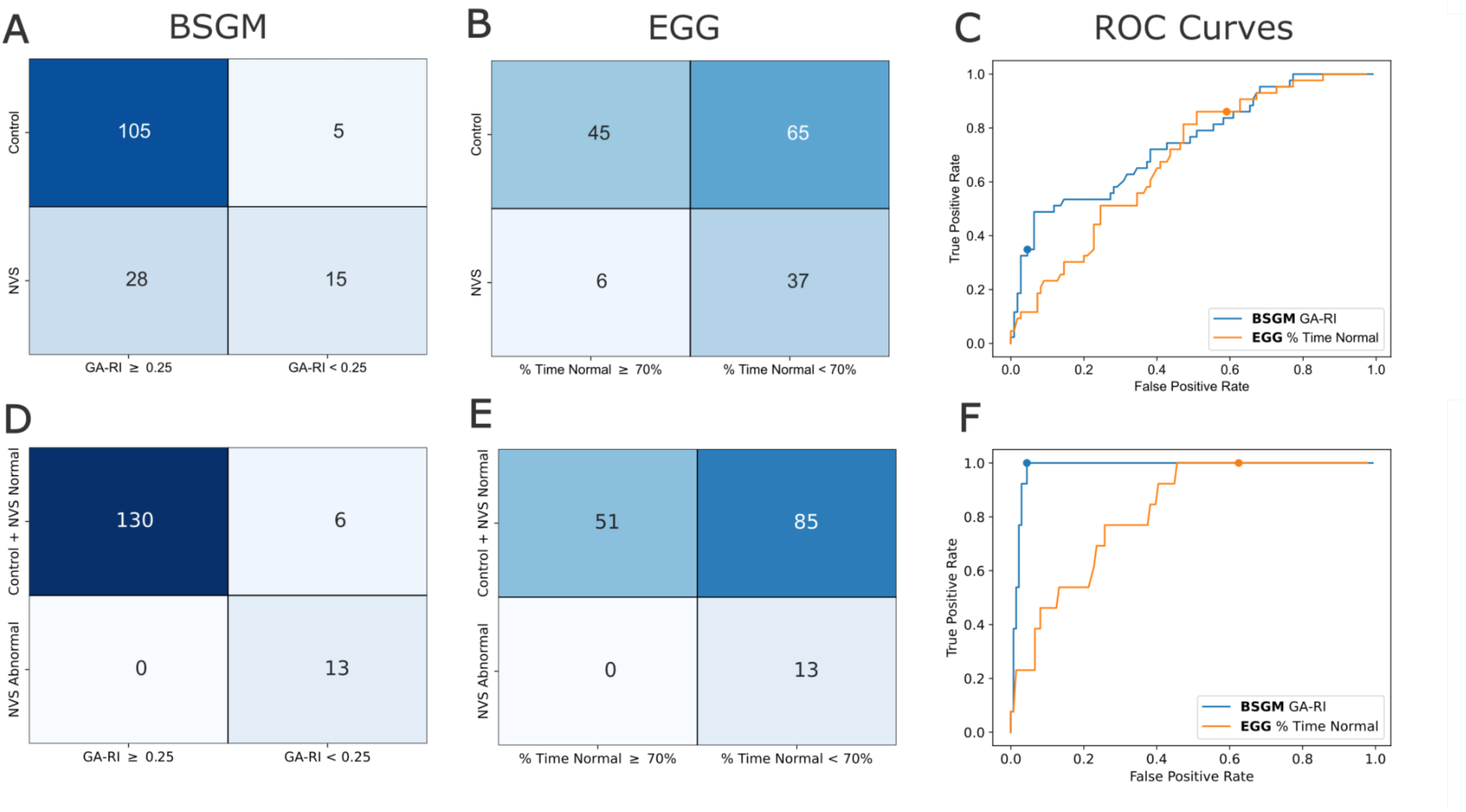
Results from individual classifications of controls vs. NVS patients **(A-C)** and control/NVS-Normal vs. NVS-Abnormal labels **(D-F)** based on rhythmic stability metrics. **(A,D)** Confusion matrix using a threshold of GA-RI ≥ 0.25 for BSGM rhythmic stability. **(B,E)** Confusion matrix using a threshold of percentage time normal frequency ≥ 70% for EGG rhythmic stability. **(C,F)** ROC curves with the location on the curve associated with the published thresholds indicated by a dot.

**Table 1.** Patient-Level Classifications of Spectral Metrics. Quantitative metrics for classification of NVS patients vs. controls and NVS-Abnomal patients vs. controls and NVS-Normal patients. Sensitivity, specificity, and accuracy were calculated using published thresholds (0.25 for BSGM and 70% for EGG), and AUC was calculated using ROC curves (i.e. with varying thresholds).

| Classification | Metric | Sensitivity | Specificity | Accuracy | AUC |
| --- | --- | --- | --- | --- | --- |
| Patient vs. Control | GA-RI (BSGM) | 0.35 | 0.95 | 0.78 | 0.65 |
|  | % Time Normal (EGG) | 0.86 | 0.41 | 0.54 | 0.50 |
| NVS-Abnormal vs. Control/ NVS-Normal | GA-RI (BSGM) | 1 | 0.96 | 0.96 | 0.82 |
|  | % Time Normal (EGG) | 1 | 0.38 | 0.43 | 0.62 |

*NVS Patient (n=43) vs. Control (n=110):* While EGG was more sensitive than BSGM (37 vs 15 out of 43 patients with low rhythm stability), it was less specific (65 vs. 5 out of 110 controls with low rhythm stability), yielding a lower overall performance for EGG than BSGM (0.54 vs 0.78 accuracy). As this tradeoff between sensitivity and specificity is affected by the threshold used for determining normal vs abnormal rhythmic stability, we varied the thresholds to produce ROC curves, finding a lower AUC for EGG than BSGM (0.50 vs. 0.65).

*NVS-Abnormal (n=13) vs. NVS-Normal/Control (n=136):* Both the BSGM and EGG pipelines had optimal sensitivity (13 out of 13 NVS-Abnormal with low rhythm stability). However, EGG misidentified 85 out of 136 subjects from the control/NVS-Normal group as having low rhythmic stability (62.5%), whereas BSGM only misidentified six (4.4%). The lack of specificity in EGG (i.e., 0.38) was not due to any single factor, but rather was representative of the collective limitations arising from all stages of the EGG pipeline, including the potential for ending the test before a delayed meal response was established, potential for missing gastric data with a single bipolar pair, lack of artifact correction, poor signal to noise ratio, and the absence of formally established normative values determined using a rigorously standardized test protocol.

To further assess the nature of the marked discrepancy in specificity between the BSGM vs EGG pipelines, average BSGM and EGG spectrograms for the controls (n=65) and NVS-Normal (n=20) individuals with low percentage time normal frequency are shown **Figure 7**. While the BSGM spectrogram is characterized by a distinct band of strong activity near 3 cpm, the EGG average spectrograms show considerable low-frequency noise and, for the NVS-Normal group, little to no activity in the gastric frequency range. This provides visual confirmation that the low specificity of the EGG-based classification is representative of its failure to reliably capture clean gastric signals in a large number of subjects with normal gastric activity. By contrast, the BSGM system was able to routinely identify a strong gastric activity band by summating activity from multiple channels overlying the stomach to obtain a high signal-to-noise ratio.

**Figure 7.**
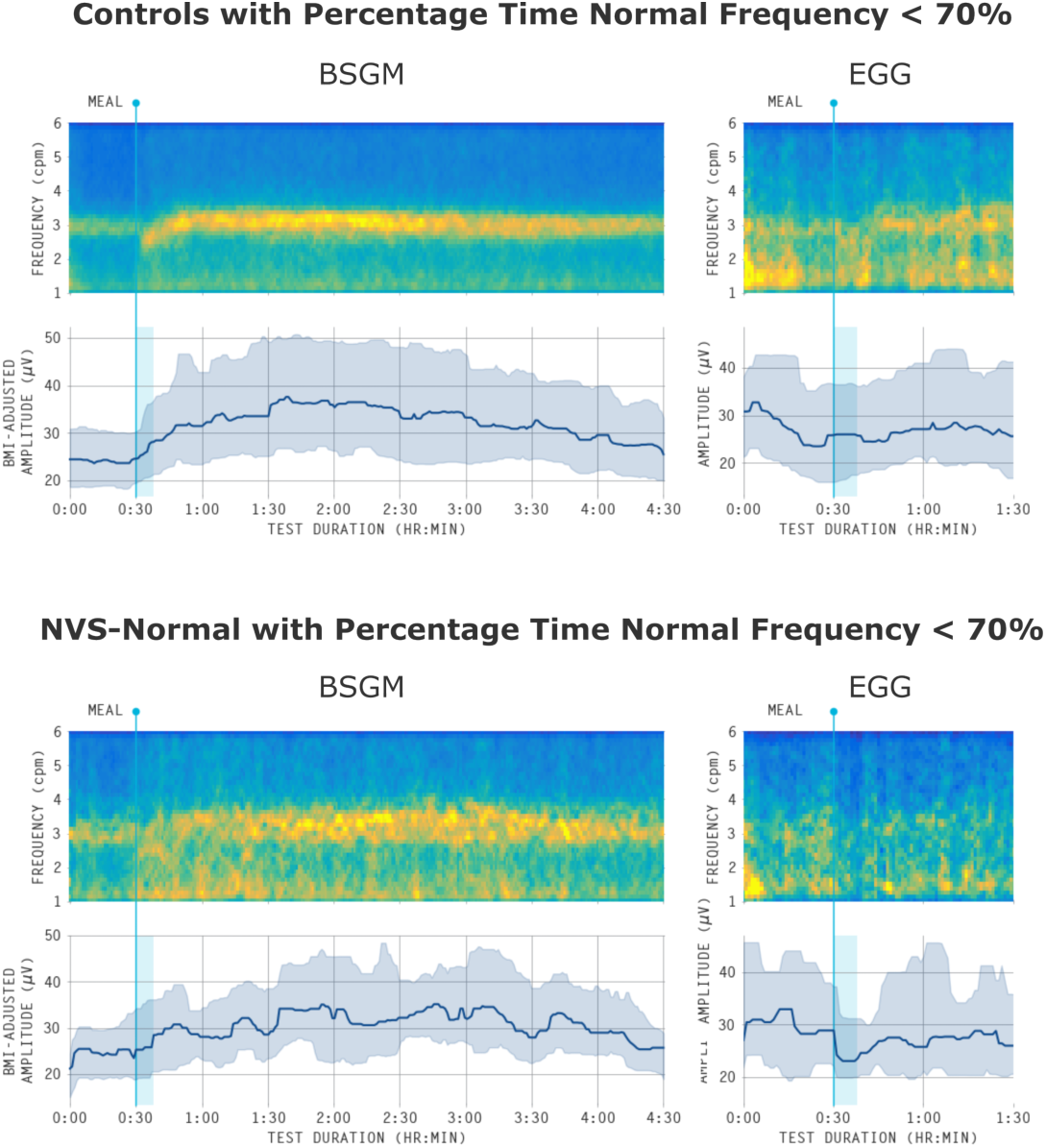
Average spectrograms for controls (top; n=65) and NVS-Normal (bottom; n=20) with percentage time normal frequency less than 70% produced by BSGM (left) and EGG (right). The lower panel in each shows the median amplitude curve with the 25th-75th percentile shaded.

## Discussion

This study presents a head-to-head comparison of Gastric Alimetry BSGM and EGG, including recording approaches and analysis pipelines. The results showed that EGG identified significant group-level differences in gastric electrophysiology between patients and controls, but lacked symptom correlations, and showed poor accuracy at patient-level classification. BSGM demonstrated marked performance improvements over EGG in all three of these domains.

EGG is now in its centennial year, and has been comprehensively evaluated in several hundred clinical studies. Systematic reviews of these studies also found that EGG consistently identifies group-level differences in gastric electrophysiology between patients and controls, including in nausea and vomiting disorders, functional dyspepsia, reflux and pediatric populations.^8–11^ Yet despite these consistent and valid findings, which provide important pathophysiological insights, EGG is widely regarded as lacking clinical utility.^3, 13, 38^ The reasons are clarified by this study. Heterogeneous mechanisms underlie symptoms in gastroduodenal disorders,^39^ and group-level differences are therefore insufficient to inform individual patient care.

EGG’s critical weakness is its poor specificity. This is emphasized by the high proportion of healthy controls with a percentage time in normal frequency below the 70% threshold. These results are consistent with the EGG literature, being within the range of a recent meta-analyses of control data.^11^ Some EGG studies reporting higher % time in normal frequency have applied non-standard techniques, e.g. filtering <1.8 cpm ^11, 40^, or used undisclosed pre-processing methods,^11, 41, 42^ such that the 70% threshold also lacks a standardized protocol for reference. We accounted for this by performing a ROC curve analysis that is independent of any singular threshold, which confirmed that the poorer performance of EGG was not simply a result of using an overly sensitive normality threshold. Moreover, the use of a 70% threshold for ‘time in normal frequency’ is non-physiological, appearing to be designed around the inherent limitations of EGG rather than derived from physiological principles, given that gastric electrophysiology mapping studies have shown that healthy control activity should lie near-continuously within the normal frequency range.^21, 22, 35, 43^

While recent literature has validated various BSGM technical advances ^16–18, 24, 44^, the present study provides the first comprehensive head-to-head comparison of the relative performance capabilities of BSGM and EGG. The essential diagnostic advantage of BSGM, as distinct from EGG, is the capability to robustly identify individual patients with or without gastric neuromuscular abnormalities, as guided by reference intervals ^22^, and as recently demonstrated in populations with NVS and T1D ^17, 23^. In addition, BSGM generates consistent symptom correlations,^17, 23, 24^ whereas EGG shows poor symptom correlations,^10, 35^ with the main finding in the literature being inconsistent associations between nausea and tachygastria.^34, 45^ It should also be recognized, however, that the consensus panel reference standard used in this study is self-referential to state-of-the-art techniques in non-invasive gastric electrophysiology, explaining the optimal sensitivity scores of 1.0 for both EGG and BSGM. A more ideal reference standard would be invasive (serosal) high-resolution gastric mapping,^46^ but this is impossible to implement at clinical scale. Nonetheless, we believe the consensus panel does serve as a fair point of comparison for EGG and BSGM. Specifically, this analysis elucidated the large number of cases for which there is clearly visible normal gastric activity using BSGM that is absent in EGG (see **Figure 7**). These discrepancies are likely attributed to a misidentification of abnormal rhythmic stability by EGG, due to limitations that affect the signal-to-noise ratio of the recorded data.^29^ Furthermore, we emphasize that BSGM yielded improved classification performance when simply comparing patients vs. controls, where the low sensitivity of BSGM can be attributed to the well-known phenomenon wherein a subset of NVS patients are expected to exhibit normal gastric activity.^17^

Spectral plots are an important output of gastric electrophysiology tests, as they provide a valuable visual aid to support quantitative test interpretation.^47^ Examples of how the Gastric Alimetry pipeline enabled markedly improved spectrogram visualizations were provided in this paper, and can be explained by three factors. First, due to a short test duration, EGG does not capture late-onset amplitude responses, which are common,^16, 21^ as per the T1D case example in **Figure 4**. Moreover, this patient’s activity showed a high Principal Gastric Frequency that was completely missed by the EGG pipeline. Secondly, Gastric Alimetry implements a highly effective validated noise cancellation algorithm,^19^ whereas EGG excludes time periods corrupted by artifact. Third, the large coverage of the BSGM array enables sophisticated denoising techniques and summation of multiple channels located directly over the stomach, providing a substantially improved signal-to-noise ratio.

Frequency profiling using the new Principal Gastric Frequency metric was also markedly more accurate than EGG dominant frequency, as explained by previous dedicated studies.^21, 22^ The reference interval for this new metric, based on 110 healthy volunteers, shows that the normal gastric frequency range is tightly defined within a normal range of 2.65 - 3.35 cpm in adults.^22^ Moreover, in a database of >1000 Gastric Alimetry cases, our collaboration has never yet observed any case to deviate from a Principal Gastric Frequency range of 2.2 - 4.1 cpm (unpublished data), a range that is also supported by invasive gastric recordings.^35^ These data highlight how bradygastria and tachygastria have been exaggerated in many past EGG studies. The conflation of frequency and rhythm metrics in EGG is also corrected in the Gastric Alimetry system, through the introduction of a distinct rhythm metric (GA-RI), a concept that was further reinforced in the current study by the finding that abnormalities in these two measures yield different symptom correlations.

There are numerous challenges to performing a fair comparison of BSGM and EGG systems. In an ideal setting, both tests would be conducted simultaneously on every subject, using dedicated hardware and processing pipelines. Unfortunately, this is technically infeasible due to the overlapping electrode setup requirements, and the undisclosed proprietary processing techniques of commercial systems. By using the same recordings in both the BSGM and EGG processing pipelines, achieved with an optimal validated electrode design,^16^ and only selecting electrodes with satisfactory impedance for the EGG analyses, we believe that the present study provides a fair comparison.

Nevertheless, there are several factors to be considered when interpreting our findings, particularly as they relate to the existing EGG literature. First, EGG analysis was conducted using an automated artifact detection algorithm. It is possible that manual approaches could have improved results, although the automated system applied here is well validated against expert manual marking.^19^ Second, proprietary signal processing steps could be used in commercial EGG devices, which are not disclosed, and hence could not be implemented here. Third, all tests were performed using a standard 482 kCal meal, and other meals or water load tests have been applied in past EGG studies; however, we selected this meal because our unpublished experience is that it generates a reliable electrophysiological response, whereas a water load does not. Fourth, we only considered a single electrode configuration. This was based on EGG literature, but multichannel EGG studies (e.g. up to 6 electrodes) have also been reported,^48, 49^ and ultrasound imaging has been applied for antral localisation.^27^ However, our simpler setup matches current commercially-available systems. Finally, all stages of the EGG processing pipeline required decisions on specific approaches for which there may not be unanimous consensus in the literature. Importantly, for all of the above considerations, our selected approach has been implemented uniformly across the entire cohort based on published guidance, with the aim of minimizing any biasing effect that could artificially impair the ability of EGG outputs to differentiate patients from controls.

Lastly, the present study focused only on spectral analyses of BSGM and EGG. While surface recordings are highly validated against invasive serosal recordings,^50^ subtle abnormalities such as conduction blocks or stable ectopic pacemakers may not be captured by non-invasive techniques applying spectral metrics.^31, 36^ Spatial analytics for non-invasive mapping are currently evolving, which could further improve the precision of BSGM and yield new phenotypes in the future.^24, 44, 51^ Moreover, use of gastric myoelectrical activity analyses for aiding in treatment and diagnosis of gastrointestinal disorders is supported by simultaneous evaluation of symptoms and complete patient medical history.

In summary, this comprehensive head-to-head comparison of Gastric Alimetry BSGM and EGG has demonstrated marked performance improvements of BSGM over EGG with regard to group-level differences, symptom correlations, and patient-level classifications. Gastric Alimetry resolves the technical limitations of EGG and presents a new non-invasive diagnostic option for gastroduodenal disorders in clinical practice.

## Data Availability

Data will be made available upon reasonable request. Requests should be made to the corresponding author, Armen Gharibans.

## Author Contributions

G.S., S.C., G.O., and A.A.G. conceived and designed the study. W.X., W.J.W., V.H., C.D.,

C.N.A., and G.O. collected the data. G.S., S.C., C.V., W.X., and A.A.G. performed data preparation and analysis. G.S., G.O., and A.A.G. drafted the manuscript. All authors reviewed and edited the manuscript.

