## Supplementary Material for "Comparison of Gastric Alimetry^®^ body surface gastric mapping versus electrogastrography spectral analysis"

Supplementary Figure S1

Average spectrograms for controls (top; n=110), T1D patients (middle; n=32), and NVS patients (bottom; n=43) for BSGM (left) and EGG (right). The lower panel in each shows the median amplitude curve with the 25th-75th percentile shaded.

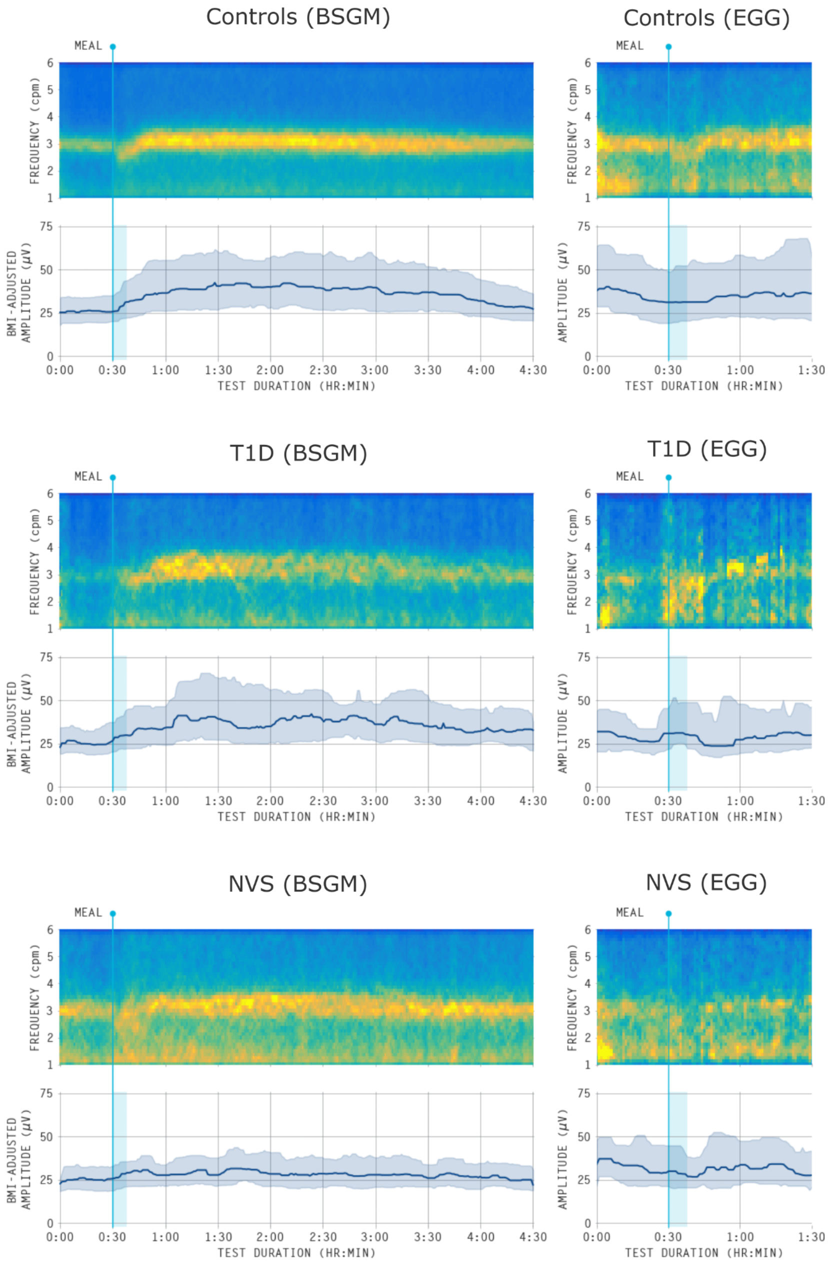

| | | Controls | T1D (v. Controls) | | | NVS (v. Controls) | | |
|---|---|---|---|---|---|---|---|---|
| | | Median (IQR) | Median (IQR) | t-statistic | p-value | Median (IQR) | t-statistic | p-value |
| BSGM Metrics | Log BMI-Adjusted Amplitude (μV) | 3.63 (3.38 - 3.93) | 3.56 (3.30 - 3.81) | 0.04 | 0.9685 | 3.38 (3.18 - 3.55) | 3.55 | **0.0005** |
| | Principal Gastric Frequency (cpm) | 3.04 (2.90 - 3.18) | 3.15 (2.97 - 3.41) | -3.63 | **0.0004** | 3.09 (2.92 - 3.27) | -1.39 | 0.1678 |
| | Gastric Alimetry Rhythm Index | 0.50 (0.39 - 0.64) | 0.43 (0.32 - 0.56) | 2.54 | **0.0121** | 0.30 (0.22 - 0.49) | 5.23 | **<0.0001** |
| | Log Fed:Fasted Amplitude Ratio | 0.62 (0.27 - 0.79) | 0.57 (0.35 - 0.81) | -0.27 | 0.7883 | 0.27 (0.09 - 0.53) | 3.61 | **0.0004** |
| EGG Metrics | Log Amplitude (μV) | 3.27 (2.82 - 3.68) | 3.04 (2.88 - 3.49) | 0.56 | 0.5747 | 3.06 (2.90 - 3.38) | 1.56 | 0.1218 |
| | Dominant Frequency (cpm) | 2.88 (1.50 - 3.12) | 2.50 (1.59 - 3.00) | 1.1 | 0.2752 | 1.62 (1.50 - 2.88) | 3.45 | **0.0007** |
| | Percentage Time Normal Frequency | 65.12 (49.05 - 82.22) | 57.85 (40.26 - 67.85) | 2.02 | **0.0453** | 48.44 (36.08 - 61.81) | 3.72 | **0.0003** |
| | Log Amplitude Ratio | 0.35 (0.10 - 0.66) | 0.34 (0.25 - 0.55) | -1.03 | 0.3065 | 0.09 (-0.06 - 0.36) | 2.96 | **0.0036** |

**Supplementary Table S1 - Group-Level Differences of Spectral Metrics**
Median and interquartile range of each metric for each participant subgroup. Results from independent t-tests are shown for each metric comparing the two patient subgroups with controls. Significant p-values (< 0.05) are shown in bold.

| Metric | Symptom | Correlation coefficient (95% CI) | p-value | Benjamini-Hochberg Critical Value |
|---|---|---|---|---|
| **Gastric Alimetry Rhythm Index** | **Bloating** | **-0.26 (-0.37 - -0.13)** | **0.0005** | **0.0018** |
| **Principal Frequency Deviation** | **Bloating** | **0.22 (0.03 - 0.40)** | **0.0034** | **0.0036** |
| **Gastric Alimetry Rhythm Index** | **Total Symptom Burden** | **-0.21 (-0.34 - -0.08)** | **0.0038** | **0.0054** |
| **Principal Frequency Deviation** | **Excessive Fullness** | **0.21 (0.04 - 0.38)** | **0.0054** | **0.0071** |
| **Gastric Alimetry Rhythm Index** | **Nausea** | **-0.20 (-0.32 - -0.08)** | **0.0062** | **0.0089** |
| **Principal Frequency Deviation** | **Upper Gut Pain** | **0.20 (0.03 - 0.39)** | **0.0074** | **0.0107** |
| **Gastric Alimetry Rhythm Index** | **Upper Gut Pain** | **-0.19 (-0.34 - -0.07)** | **0.0087** | **0.0125** |
| **% Time Normal Frequency** | **Bloating** | **-0.18 (-0.31 - -0.04)** | **0.013** | **0.0143** |
| **Principal Frequency Deviation** | **Total Symptom Burden** | **0.18 (0.03 - 0.35)** | **0.0148** | **0.0161** |
| Gastric Alimetry Rhythm Index | Excessive Fullness | -0.16 (-0.29 - -0.02) | 0.0325 | 0.0179 |
| Dominant Frequency Deviation | Bloating | 0.15 (0.01 - 0.28) | 0.04 | 0.0196 |
| BMI-Adjusted Amplitude | Heartburn | 0.13 (-0.13 - 0.35) | 0.0712 | 0.0214 |
| % Time Normal Frequency | Total Symptom Burden | -0.13 (-0.27 - 0.02) | 0.0718 | 0.0232 |
| Dominant Frequency Deviation | Total Symptom Burden | 0.13 (-0.00 - 0.26) | 0.0787 | 0.025 |
| Principal Frequency Deviation | Nausea | 0.13 (0.00 - 0.28) | 0.0794 | 0.0268 |
| Fed:Fasted Amplitude Ratio | Bloating | -0.13 (-0.26 - 0.06) | 0.0871 | 0.0286 |
| % Time Normal Frequency | Excessive Fullness | -0.12 (-0.27 - 0.03) | 0.0928 | 0.0304 |
| Dominant Frequency Deviation | Nausea | 0.11 (-0.03 - 0.25) | 0.1403 | 0.0321 |
| Dominant Frequency Deviation | Excessive Fullness | 0.11 (-0.03 - 0.24) | 0.152 | 0.0339 |
| Gastric Alimetry Rhythm Index | Stomach Burn | -0.10 (-0.24 - 0.03) | 0.1677 | 0.0357 |
| % Time Normal Frequency | Nausea | -0.10 (-0.24 - 0.04) | 0.1891 | 0.0375 |
| Dominant Frequency Deviation | Upper Gut Pain | 0.10 (-0.05 - 0.24) | 0.1902 | 0.0393 |
| Fed:Fasted Amplitude Ratio | Total Symptom Burden | -0.09 (-0.25 - 0.09) | 0.215 | 0.0411 |
| BMI-Adjusted Amplitude | Nausea | -0.08 (-0.22 - 0.07) | 0.2704 | 0.0429 |
| % Time Normal Frequency | Upper Gut Pain | -0.08 (-0.22 - 0.07) | 0.2864 | 0.0446 |
| Fed:Fasted Amplitude Ratio | Nausea | -0.08 (-0.25 - 0.09) | 0.2995 | 0.0464 |
| Amplitude | Bloating | -0.08 (-0.17 - 0.05) | 0.3072 | 0.0482 |
| Fed:Fasted Amplitude Ratio | Upper Gut Pain | -0.07 (-0.26 - 0.15) | 0.313 | 0.05 |
| Dominant Frequency Deviation | Heartburn | 0.07 (-0.07 - 0.24) | 0.3464 | 0.0518 |
| % Time Normal Frequency | Stomach Burn | -0.07 (-0.24 - 0.11) | 0.3506 | 0.0536 |
| Dominant Frequency Deviation | Stomach Burn | 0.07 (-0.07 - 0.21) | 0.3517 | 0.0554 |
| Amplitude Ratio | Bloating | -0.07 (-0.23 - 0.12) | 0.3784 | 0.0571 |

| | | | | |
|---|---|---|---|---|
| Gastric Alimetry Rhythm Index | Heartburn | -0.06 (-0.24 - 0.08) | 0.3954 | 0.0589 |
| Amplitude Ratio | Stomach Burn | 0.06 (-0.16 - 0.35) | 0.4253 | 0.0607 |
| Amplitude Ratio | Total Symptom Burden | -0.05 (-0.24 - 0.15) | 0.4601 | 0.0625 |
| Amplitude Ratio | Heartburn | 0.05 (-0.11 - 0.23) | 0.4721 | 0.0643 |
| BMI-Adjusted Amplitude | Bloating | -0.05 (-0.19 - 0.10) | 0.5069 | 0.0661 |
| Fed:Fasted Amplitude Ratio | Excessive Fullness | -0.05 (-0.20 - 0.13) | 0.5181 | 0.0679 |
| Principal Frequency Deviation | Heartburn | -0.04 (-0.14 - 0.09) | 0.5919 | 0.0696 |
| BMI-Adjusted Amplitude | Total Symptom Burden | -0.04 (-0.19 - 0.16) | 0.6089 | 0.0714 |
| Amplitude | Heartburn | 0.04 (-0.08 - 0.19) | 0.6317 | 0.0732 |
| Fed:Fasted Amplitude Ratio | Stomach Burn | 0.03 (-0.18 - 0.27) | 0.6615 | 0.075 |
| Principal Frequency Deviation | Stomach Burn | 0.03 (-0.12 - 0.18) | 0.6719 | 0.0768 |
| Fed:Fasted Amplitude Ratio | Heartburn | 0.03 (-0.14 - 0.21) | 0.7308 | 0.0786 |
| Amplitude | Stomach Burn | 0.02 (-0.11 - 0.19) | 0.7397 | 0.0804 |
| Amplitude Ratio | Upper Gut Pain | -0.02 (-0.24 - 0.24) | 0.7499 | 0.0821 |
| Amplitude Ratio | Nausea | -0.02 (-0.22 - 0.17) | 0.7612 | 0.0839 |
| % Time Normal Frequency | Heartburn | -0.02 (-0.19 - 0.13) | 0.8114 | 0.0857 |
| Amplitude | Total Symptom Burden | -0.02 (-0.13 - 0.12) | 0.8179 | 0.0875 |
| Amplitude | Excessive Fullness | 0.02 (-0.11 - 0.17) | 0.8359 | 0.0893 |
| Amplitude Ratio | Excessive Fullness | -0.01 (-0.20 - 0.19) | 0.8442 | 0.0911 |
| BMI-Adjusted Amplitude | Upper Gut Pain | -0.01 (-0.20 - 0.17) | 0.8557 | 0.0929 |
| BMI-Adjusted Amplitude | Stomach Burn | 0.01 (-0.19 - 0.23) | 0.8734 | 0.0946 |
| Amplitude | Nausea | -0.01 (-0.13 - 0.12) | 0.8749 | 0.0964 |
| Amplitude | Upper Gut Pain | -0.01 (-0.13 - 0.14) | 0.9035 | 0.0982 |
| BMI-Adjusted Amplitude | Excessive Fullness | -0.01 (-0.15 - 0.16) | 0.9203 | 0.1 |

**Supplementary Table S2 - Correlation of Spectral Metrics with Symptoms**

Complete results of the metric/symptom correlation analysis. For each metric/symptom pair, we reported the Pearson correlation coefficient with 95% confidence intervals computed using a bootstrapping procedure with 1000 bootstrap samples, the associated p-value, and Benjamini-Hochberg critical values. Data is ordered by p-value and bolded for rows that are significant according to the Benjamini-Hochberg procedure
